# Interpreting Mendelian randomization estimates of the effects of categorical exposures such as disease status and educational attainment

**DOI:** 10.1101/2020.12.14.20248168

**Authors:** Laurence J Howe, Matthew Tudball, George Davey Smith, Neil M Davies

## Abstract

Mendelian randomization has been previously used to estimate the effects of binary and ordinal categorical exposures - e.g. type 2 diabetes or educational attainment defined by qualification - on outcomes. Binary and categorical phenotypes can be modelled in terms of liability, an underlying latent continuous variable with liability thresholds separating individuals into categories. Genetic variants typically influence an individual’s categorical exposure via their effects on liability, thus Mendelian randomization analyses with categorical exposures will capture effects of liability which act independent of exposure category.

We discuss how groups where the categorical exposure is invariant can be used to detect liability effects acting independently of exposure category. For example, associations between an adult educational attainment polygenic score (PGS) and BMI measured before the minimum school leaving age (e.g. age 10), cannot indicate the effects of years in full-time education on this outcome. Using UK Biobank data, we show that a higher education PGS is strongly associated with lower smoking initiation and higher glasses use at age 15. These associations were replicated in sibling models. An orthogonal approach using the raising of the school leaving age (ROSLA) policy change found that individuals who chose to remain in education to age 16 before the reform likely had higher liability to educational attainment than those who were compelled to remain in education to 16 after the reform, and had higher income, decreased cigarette smoking, higher glasses use and lower deprivation in adulthood. These results suggest that liability to educational attainment associates with health and social outcomes independent of years in full-time education.

Mendelian randomization studies with non-continuous exposures should be interpreted in terms of liability, which may affect the outcome via changes in exposure category and/or independently.

## Introduction

Mendelian randomization can be implemented as an instrumental variables analysis using genetic variants to evaluate causal relationships of potential exposures (e.g. LDL-cholesterol) on outcomes (e.g. coronary heart disease) ^1,2^. The Wald estimator – the ratio of the associations of the genetic variants with the outcome and the exposure is an estimator of the effect of an exposure on an outcome. Mendelian randomization analyses require the three core instrumental variable assumptions; 1) the genetic variants are associated with the exposure (relevance), 2) there are no unmeasured confounders of the genetic variant-outcome association (independence) and 3) that the genetic variants only influence the outcome via their effect on the exposure (the exclusion restriction) ^3-5^. Mendelian randomization has been widely used to estimate the effects of continuous exposures and to estimate the effects of binary and ordinal categorical exposures such as type II diabetes status ^6-9^ and educational attainment ^10-12^.

Conceptually, binary exposures, such as disease status, can be modelled by assuming an underlying continuous liability, a normally-distributed latent (unmeasured) variable ^13-20^. Liability models can be deterministic as in the Falconer liability-threshold model ^13,16^, where an individual’s binary disease status is completely determined by whether their underlying continuous liability to disease exceeds a threshold. Here, liability is a combined measure of all sources of variation influencing disease risk; genetic variation, the environment and chance ^13,15,21^. Although the liability-threshold model is most often used for binary disease states, it can be extended to ordinal categorical phenotypes ^22^, i.e. any phenotype with a finite number of possible values but a clear continuous ordered dimension. For example, considering years spent in full-time education, individuals across a population have an underlying liability to educational attainment and their duration in full-time education category is determined by their liability with respect to multiple population-level thresholds **(Figure 1)**.

**Figure 1.**
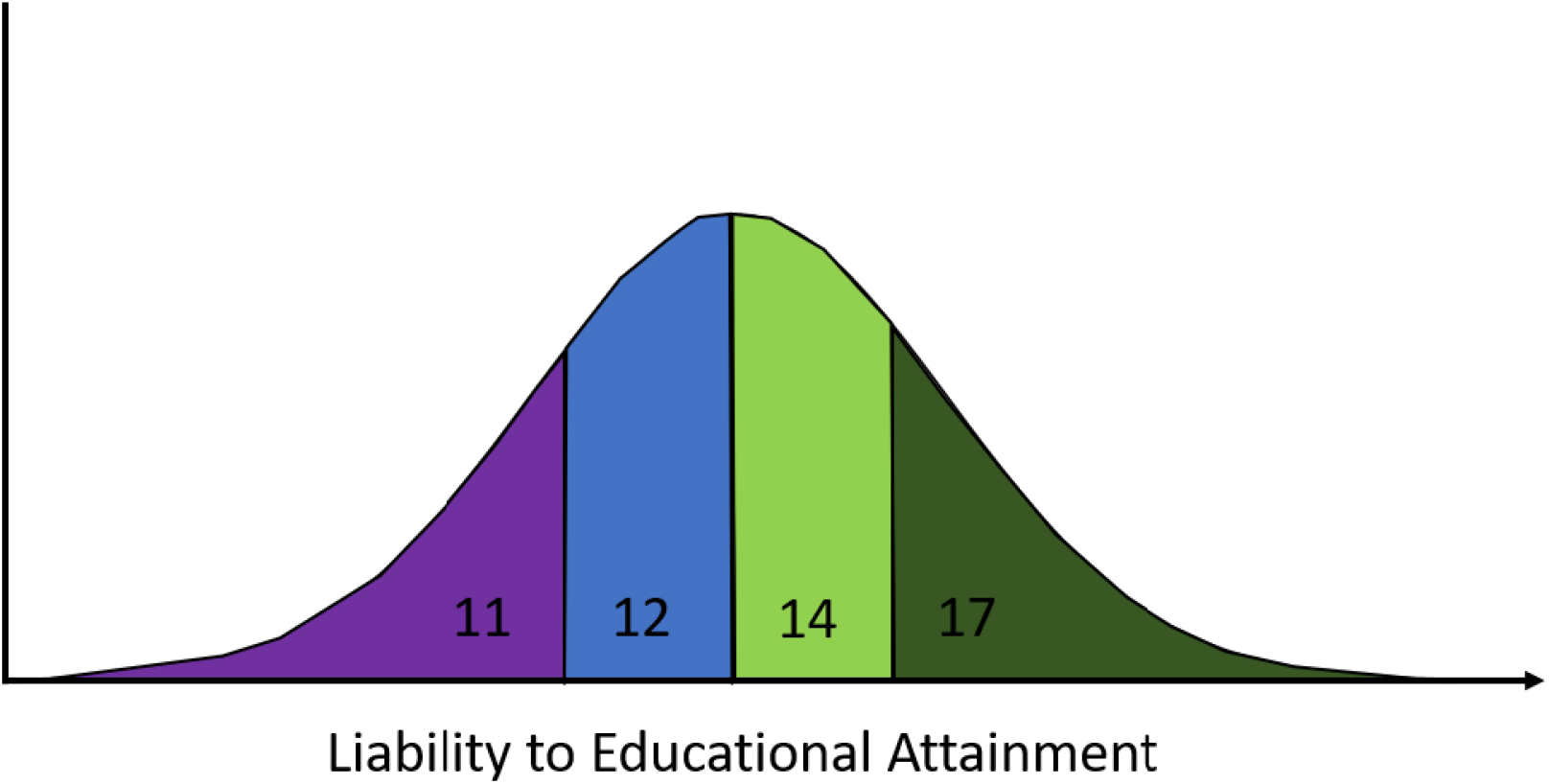
Years spent in full-time educational attainment as a function of underlying continuous liability. Individuals in the population can spend 11, 12, 14 or 17 years in full-time education (started at the age of 4 years and left aged 15, 16, 18 and 21 respectively). Educational attainment category is determined by their underlying liability with respect to population-level thresholds.

Here, we start by discussing how to interpret effects of categorical exposures in Mendelian randomization analyses. In particular, we describe three possible causal mechanisms between a categorical exposure and an outcome. Next, we outline how performing analyses in a population where the categorical exposure is invariant can be used to inform the most likely causal mechanism. We then use empirical data from UK Biobank to evaluate if educational attainment polygenic scores (PGS), used as proxies for liability to years in schooling, are associated with childhood phenotypes (glasses wearing, smoking initiation and body mass index). As the outcomes are measured in children below the minimum school leaving age, associations cannot reflect effects of years in schooling. Finally, we use a previous UK-wide school reform and UK Biobank data to further investigate the mechanism via which education affects later-life outcomes.

### Theoretical basis for estimating effects of categorical exposures using Mendelian randomization

The interpretation of non-continuous exposures in Mendelian randomization analyses has been previously discussed ^23-25^. Taylor et al. (2014) described how Mendelian randomization analyses of the effects of smoking using categorical exposures based on reported cigarettes per day, are likely to be biased compared to more precise measures such as blood cotinine ^23^. Burgess and Labrecque (2018) discussed how to interpret the effects of binary exposures. In particular, they note that the exclusion restriction assumption requires that the genetic variants do not influence the outcome except via the binary exposure status ^25^. For example, genetic variants may only influence chemotherapy treatment status via cancer diagnosis. If this does not hold, for example if the underlying latent continuous trait influences the outcome independent of the binary exposure, then this would lead to bias ^25,26^. We extend previous discussion by outlining theoretical models for interpreting the effects of categorical (binary or ordinal) exposures.

Generally, Mendelian randomization studies with binary or ordinal categorical exposures are interested in estimating the effect of a change in category (*C*). For example, estimating the effect of being exposed or not for a binary exposure (e.g. the effect of having a disease or not), or the effect of increasing by a level of categorical variable (e.g. the effect of getting 12 rather than 11 years of education) – rather than effects of the underlying liability to the categorical variable (*L*) ^24,27^. However, genetic instruments, with the possible exception of some variants relating to monogenic phenotypes, are not deterministic and so influence *L*.

In many instances, *L* could plausibly affect an outcome *O* via pathways involving changes in *C* and also via pathways independent of *C* (*l*) (equations 1/2). For example, liability to myocardial infarction (MI) could clearly impact mortality via having an MI event but underlying liability to MI relating to atherosclerotic disease could also affect mortality amongst individuals independent to having an MI event. Similarly, a combination of type 2 diabetes and related metabolic disturbances may contribute to the increased incidence of coronary heart disease risk amongst type 2 diabetics ^28,29^. It follows that the association between genetic instruments for a categorical exposure and *O* will capture effects of *L* on *O*, via both *l* and *C*. If the effects via *l* are non-zero then this would lead to a violation of the exclusion-restriction assumption in a Mendelian randomization analysis.

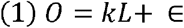

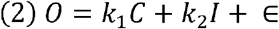

*k* = total effect of *L* on *O, k*_l_ = effect of *L* on *O* mediated by *C, k*_2_ = effect of *L* on *O* mediated by *l*, ∈= remaining sources of variation in *O*

Three possible causal mechanisms between a categorical exposure and an outcome are:

A. the liability-exclusive model: *L* influences *O* but via pathways *l*, independent of *C*;
B. the threshold-exclusive model: *L* influences *O* only via threshold effects relating to changes in *C* (stepwise);
C. the combined model: *L* influences *O* both via *l* and *C*.

Mendelian randomization estimates can be interpreted as reflecting each of these models under different assumptions. For the estimates to reflect the liability exclusive model (A above), we must assume that changing *C*, holding *L* constant, has no effect on the outcome. For example, a Mendelian randomization study of obesity may suggest that obesity affects risk of type II diabetes. However, this relationship is likely entirely driven by the underlying variable BMI. Alternatively, we could assume that the estimates solely reflect the effect of the threshold model (B above) and that *L* has no effect on the outcome except via *C*. For example, children born with orofacial clefts require surgery, this is likely to be entirely mediated by whether someone has a cleft or not, with the underlying liability having no impact independent of cleft status (**Figure 2**). Finally, it is possible that the estimates reflect a combination of the two models, i.e. effects of *L* mediated by, and independent of, *C*.

**Figure 2.**
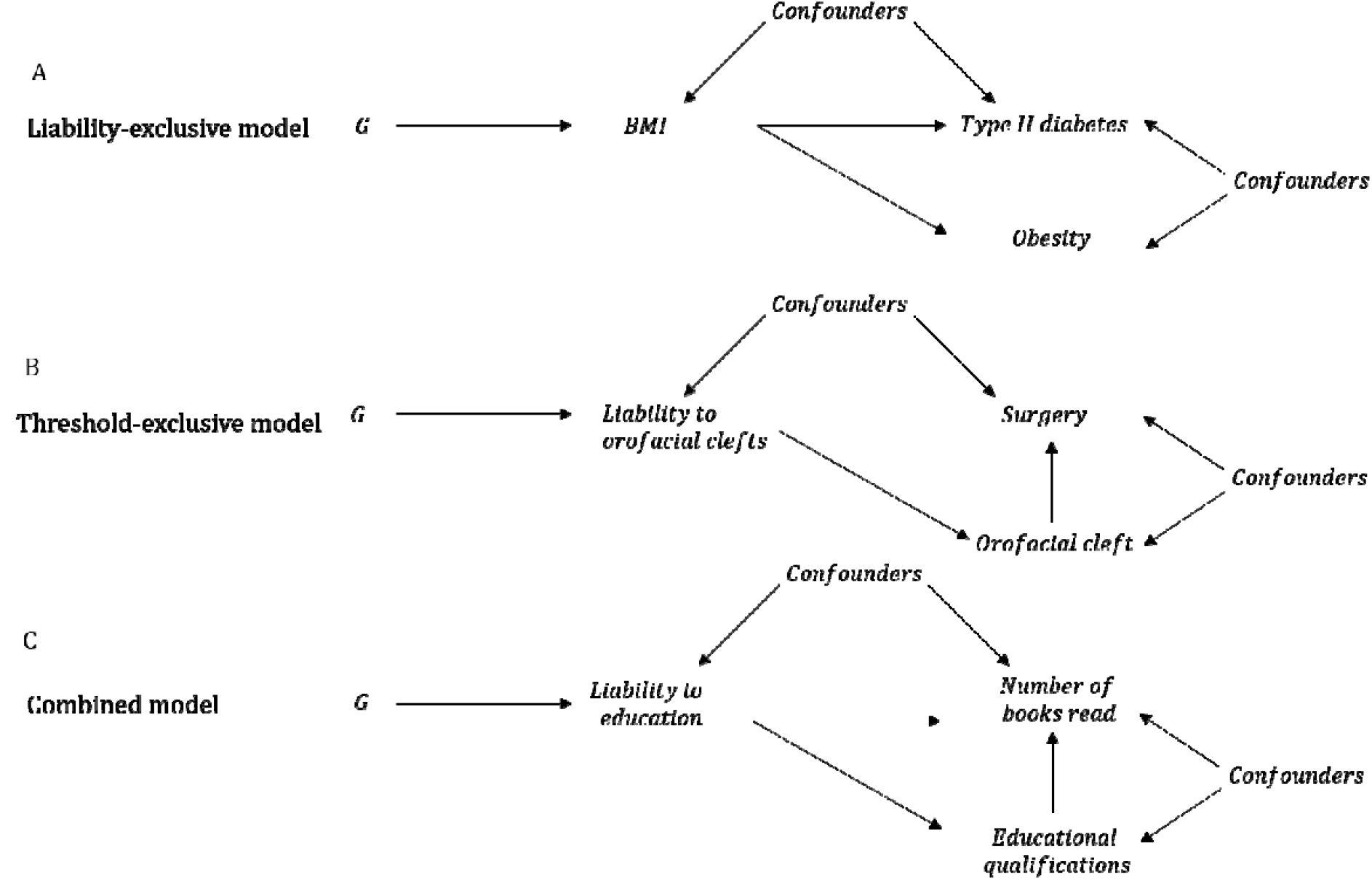
Causal graph illustrations of interpreting causal relationships between non-continuous exposures and outcomes. Illustrations of the liability-exclusive, threshold-exclusive and combined models for interpreting causal relationships between ordinal categorical exposures and outcomes. G=genetic factors, BMI=body mass index.

A. Under the liability-exclusive model, liability influences the outcome solely via effects which are independent of exposure category. For example, an MR study of obesity (i.e. BMI>30) and type II diabetes would suggest that obesity increases risk of type II diabetes risk. However, this effect is likely to be solely due to the effects of continuous body mass index (liability to obesity) rather than threshold effects relating to body mass index categories.
B. In the threshold-exclusive model, liability to the exposure influences the outcome entirely via threshold effects relating to the categories of the exposure (i.e. a stepwise effect). For example, individuals born with an orofacial cleft are likely to have corrective surgery but individuals who do not develop an orofacial cleft will not, irrespective of their underlying liability to orofacial clefts.
C. In the combined model, liability influences the outcome via the categorical exposure and via pathways independent of the categorical exposure. For example, spending longer in full-time education involves reading books but individuals with high liability to educational attainment may also be more likely to read books independent of educational attainment.

### Evaluating the “threshold-exclusive” model

The “threshold-exclusive” model can be tested using genetic variants associated with the exposure in a population where there is no variation in the categorical exposure. Consider the example of genetic risk variants associated with coronary heart disease in adults amongst children, a group likely to be free of coronary heart disease ^30^. In this context, assuming no confounders of the genetic instrument-outcome association (independence) and that the genetic components of liability have the same effect on the outcome as environmental components (gene-environmental equivalence), associations between a coronary heart disease genetic score and outcomes (e.g. metabolites in childhood ^28,30^) would be indicative of effects of liability to coronary heart disease independent of disease status.

Another approach could be to stratify individuals by exposure category, i.e. split a study of elderly adults into individuals with and without coronary heart disease and determine if the genetic score is associated with the outcome within each stratum. This approach is not generally advisable because stratifying on the exposure will likely induce collider bias. For example, disease-free individuals with high genetic risk for coronary heart disease are likely to have low non-genetic liability to coronary disease ^31^ (**Figure 3**). Similarly, measurement error could induce associations in the stratified analysis even if effects are entirely via threshold effects.

**Figure 3.**
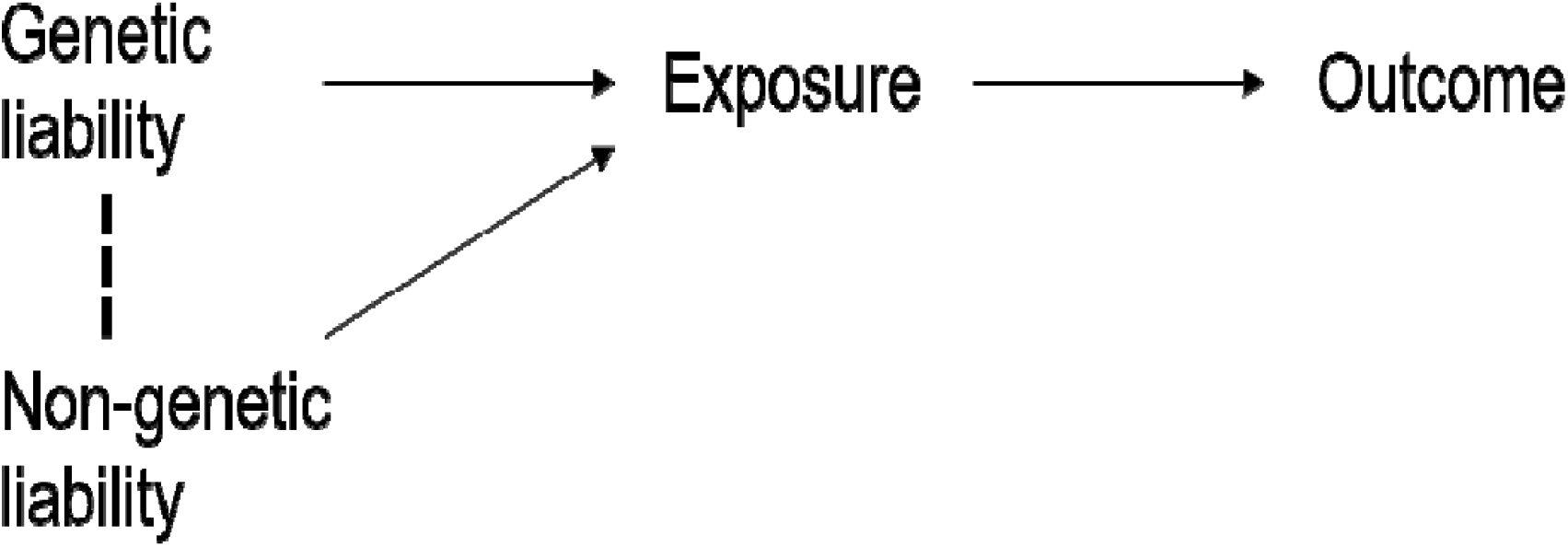
A causal graph illustrating potential for associations between genetic and non-genetic liability when conditioning on exposure category. Stratifying on exposure category, associations could be induced between genetic and non-genetic determinants of liability. For example, if a diseased case has low genetic liability to a disease then dependent on the model, they may be more likely to have higher non-genetic liability (environment/stochastic).

Analysis in stratified subgroups can be susceptible to collider bias but stratification on factors that cannot be influenced by the exposure and the outcome are unlikely to induce bias (e.g. sex or age) ^32,33^. If an exposure is invariant in either sex, in an age group or perhaps in a country, it is possible to evaluate the “threshold-exclusive” model without collider concerns. For example, a similar approach has been used in previous Mendelian randomization studies of alcohol consumption in East Asian populations to investigate whether associations between alcohol-related genetic variants and outcomes are via alcohol consumption. In these populations, women have very low alcohol consumption compared to men so if the effect is via alcohol consumption then associations between genetic variants and outcome should be null in women (assuming they consume very low to zero alcohol) ^34-36^.

### Educational attainment PGS and childhood phenotypes in UK Biobank

We sought to use a population invariant for educational attainment (as measured by years in schooling) and an educational attainment polygenic score (PGS) to investigate the “threshold-exclusive model”. We used self-reported childhood phenotype data from adults in UK Biobank; relative body size at age 10, wearing glasses by age 15 and smoking initiation by age 15. The minimum school leaving age during this time-period was 15 (or 16 from 1972 onwards) so associations between the education PGS and these childhood phenotypes cannot relate to effects of additional schooling (), but could relate to effects of liability to education ()

Using a sample of 337,006 individuals of recent European ancestry, we found strong evidence that a 1 SD higher education PGS (173 independent variants with P < 1×10^−15^ from an independent sample) is associated with higher odds of wearing glasses (OR 1.05; 95% C.I. 1.04, 1.06) and lower odds of smoking initiation (OR 0.88; 95% C.I. 0.87, 0.89), both at age 15. Contrastingly, we did not find strong evidence for an association between the education PGS and body-size at age 10 (per SD increase in PGS: Beta = 0.000; 95% C.I. −0.002, 0.003), although the measure was ordinal rather than continuous, limiting power (**Table 1**). These results suggest that *E*^*^ increases the wearing of glasses and reduces smoking initiation independent of *E*.

**Table 1.** Education PGS and pre-adulthood BMI, smoking and glasses use

| Outcome |  | Change per 1 S.D. increase in Education PGS (95% C.I.) |
| --- | --- | --- |
| Body-size at age 10: (0, 1, 2)* | Beta (95% C.I.) | 0.000 (-0.002, 0.003) |
| Wear glasses at age 15: (yes/no) | OR (95% C.I.) | 1.05 (1.04, 1.06) |
| Smoking at age 15: (yes/no) | OR (95% C.I.) | 0.88 (0.87, 0.89) |

\*Comparative body size at age 10 to peers; 0 = Thinner, 1 = Average, Plump = 2

To investigate if familial factors (e.g. effects of parental education on offspring phenotypes^37^) were driving these associations, we repeated analyses for glasses wearing and smoking phenotypes using a within-family model in UK Biobank. Using a sample of 41,497 siblings from 19,588 families, we found consistent evidence that the educational attainment PGS was associated with increased glasses wearing (per SD increase in PGS; OR 1.05; 95% C.I. 1.01, 1.10) and reduced smoking initiation (per SD increase in PGS; OR 0.89; 95% C.I. 0.84, 0.94) at age 15. These results are further indicative of effects of *E*^*^ independent of *E*.

### Raising of the school age and liability to education

An alternative, and widely used, source of evidence about the effects of education are policy reforms which affected the amount of schooling people received – such as the raising of the school leaving age (ROSLA). In 1972 the minimum school leaving was increased by one year from 15 to 16 years old. This meant that individuals born before September 1957 could leave at age 15, where those born in September 1957 or afterwards had to stay in school until at least age 16. ROSLA has been previously used to estimate the effects of an additional year in school on health outcomes in UK Biobank ^27^.

Here, we use the schooling reform to investigate whether individuals who would have chosen to stay in school to 16 even before the reform, have better outcomes than individuals who would prefer to leave, this is a measure of *E*^*^. To distinguish this analysis from the ROSLA analyses described above, which investigate the effects of additional education, we refer to this analysis as ROSLA-L. We selected participants who reported leaving school at age 16 in the year before the reform (born September 1956 – August 1957) and the year after the reform (born September 1957 – August – 1958). This sample consists of individuals who were not affected by the reform (pre-reform cohort), who chose to remain in school until age 16, and those who were affected by the reform and may have been forced to stay until age 16 (post-reform cohort). The post-reform cohort will include individuals who would have preferred to leave school at age 15 given the choice. We estimated the difference in outcomes between the pre- and post-reform cohorts. Assuming no time effects, differences in outcomes between the pre- and post-reform cohorts are likely to reflect group-level differences between individuals choosing to stay in school until 16 and individuals who would have preferred to leave at 15.

In UK Biobank, there were 2,592 participants in who left school at age 16 in the pre-reform cohort and 4,064 who left school at 16 in the post-reform cohort from England and Wales. Individuals in the post-reform cohort had lower average education PGS (−0.06 SD; 95% C.I. −0.11, −0.01) than the pre-reform cohort, indicative of differences in *E*^*^ as hypothesised. We found strong evidence that the post-reform cohort had higher pack years of smoking, a lower proportion with household income over £18,000 and higher deprivation than the pre-reform cohort. Contrastingly, there was little evidence for group-level differences in BMI or SBP, but conclusions are limited by modest sample sizes (**Table 2**).

**Table 2.** Adulthood phenotypic differences between the pre- and post- reform cohorts

| Phenotype | Pre-reform (N = 2,592)<br>Left school at 16 year<br>before reform | Post-reform (N = 4,064)<br>Left school at 16 year<br>after reform | Heterogeneity P-value |
| --- | --- | --- | --- |
| BMI: mean (SD) | 27.9 (5.0) | 28.0 (5.0) | 0.40 |
| Smoking pack years:<br>mean (SD) | 21.8 (15.3) | 23.7 (17.1) | 0.0095 |
| SBP: mean (SD) | 136.6 (18.0) | 136.2 (18.0) | 0.35 |
| Townsend deprivation<br>index: mean (SD)<sup>1</sup> | -1.33 (3.0) | -0.93 (3.1) | 2.6×10<sup>-7</sup> |
| Household income:<br>(% with over £18,000) | 83.2% | 80.5% | 0.0080 |
| Glasses wearing<sup>2</sup>:<br>(yes/no) | 90.2% | 88.0% | 0.0051 |

<sup>1</sup> a greater score implies a greater degree of deprivation
<sup>2</sup> includes contact lenses

The ROSLA-L results suggest that individuals with higher *E* ^*^ smoke less, have lower deprivation, have higher income and are more likely to wear glasses independent of *E*. However, we cannot determine whether these associations relate to direct effects of *E*^*^ or reflect effects of one component of liability (e.g. effects of parental education) but not all of the components (which would be effects of *E*^*^).

## Discussion

Here, we discuss how to interpret Mendelian randomization studies with non-continuous (binary or ordinal categorical) exposures, extending previous literature on binary exposures ^24,25,27^. Phenotypic variation in binary disease outcomes is often modelled using liability ^13^, an underlying latent continuous trait reflecting genetic, environmental and stochastic components, and we describe how the liability model can also be applied to ordinal categorical traits. Interpretation of effects of non-continuous exposures from Mendelian randomization analyses is nuanced because genetic instruments for categorical exposures capture liability to the categorical exposure rather than purely variation in the categorical exposure itself. Interpreting effect estimates in terms of the categorical exposure requires making assumptions about how genetic instruments influence the outcome, either by their effect on the exposure category (“threshold-exclusive” model), their effect on the underlying liability (“liability-exclusive” model) or some combination of both (“combined” model).

We discussed how this assumption can potentially be evaluated by determining whether genetic instruments are associated with the outcome in subgroups where the exposure category is invariant. For example, selecting a subgroup of the population who cannot have the categorical exposure, i.e. testicular cancer in women. We recommend only stratifying on factors which cannot be influenced by the exposure and outcome (e.g. sex or age) because stratification on other factors could induce collider bias, complicating interpretation. Similar negative control approaches have been used previously in Mendelian randomization analyses as sensitivity analyses to test if genetic instruments are associated with the outcome when the exposure is invariant. For example, sex-stratified analyses have been used in the context of alcohol consumption in East Asian populations exploiting alcohol behaviour differences between men and women ^34-36^. Previous studies have also looked at associations between PGS for adulthood diseases and cigarette smoking and outcomes in cohorts of children ^30,38,39^. Here, associations are likely to relate to effects of disease-liability because children are unlikely to have experienced disease events or started smoking (if under a certain age).

For educational attainment, one could similarly perform analysis using phenotypes from individuals under the minimum school leaving age, as effects of the genetic instruments in this population cannot be via additional schooling. For example, previous studies have showed that educational attainment PGS are associated with childhood school performance suggestive of liability-exclusive effects ^40-42^. Using this approach, we found evidence that educational attainment PGS are associated with glasses use and smoking initiation by age 15 as recalled by adult UK Biobank participants. These associations were replicated in within-family models suggesting that the associations were not driven by indirect genetic effects, assortative mating or population stratification ^43-45^. One limitation of this analysis is that the childhood phenotypes were recalled by adult study participants decades later so are susceptible to measurement error. An orthogonal approach, ROSLA-L, using a policy change also provided evidence that individuals with higher liability to education have better later-life outcomes independent of schooling. The PGS and ROSLA-L results indicate that liability to education likely effects health and social outcomes independent of educational thresholds. For example, individuals who are more likely to enrol in additional education may smoke less independent of education.

This has implications for Mendelian randomization analyses of the effects of educational attainment; genetic variants are proxying for liability to education so causal estimates are unlikely to reflect pure effects of additional education. Indeed, scaling Mendelian randomization estimates in terms of years of schooling may incorrectly attribute liability-exclusive effects to effects of additional schooling. Mendelian randomization has been previously used to demonstrate that measured educational attainment influences myopia and smoking behaviour ^46 47^. The associations we report here between the education PGS and related outcomes in 15-year olds, suggest that the reported effects of additional schooling on these outcomes are likely to be overestimated.

In general, genetic instruments are unlikely to only affect an outcome via changes in exposure category but may be the case when an intervention relates to a specific threshold. For example, being prescribed statins may relate to cholesterol levels reaching a certain threshold or being born with a birth defect (e.g. an orofacial cleft) could result in disrupted schooling because of surgical interventions ^48^. Mendelian randomization analyses with ordinal categorical exposures can be interpreted in terms of effects of liability which act via and independently to the categorical exposure. For example, our findings suggest that Mendelian randomization analyses of educational attainment should be interpreted as effects of liability to education which are likely to be mediated by a combination of measured educational attainment and other independent factors. We note that conventional Mendelian randomization which divides by the genetic variant-categorical exposure association will provide biased estimates of the total effect of liability because there are pleiotropic pathways from the genetic variant to the outcome within levels of the categorical exposure. With some additional steps, the total effect of liability can be estimated assuming a liability-threshold model for the genetic variant-categorical exposure relationship ^49^. In principle, future work could combine prospective data and dated events with Mendelian randomization to disentangle threshold and liability-exclusive effects as such investigation is limited with existing datasets.

In comparison to Mendelian randomization, ROSLA analyses ^27,50-52^ will identify effects of continuous education (i.e. days in the classroom), and also the effects of getting any qualifications (the threshold). ROSLA estimates using the entire sample, will not capture effects of individual liability to education, as population level average liability to education is unlikely to have changed before and after the reform. However, the average liability to education of those who chose to remain in school to 16 before the reform, is likely to be higher than those who remain in school to age 16 after the reform, as the latter includes individuals who would otherwise have left at age 15.

To conclude, we have demonstrated that interpreting Mendelian randomization with non-continuous categorical exposures requires assumptions about the underlying causal model. We described how this assumption could be evaluated using subsets of the population invariant for the exposure. However, the practical uses of this test are limited because stratifying can induce collider bias unless the stratifying factor cannot be influenced by the exposure or outcome. Evidence from genetic and school reform analyses suggested that liability to education affects social and health outcomes in UK Biobank independent of categorical measures of education attainment. Mendelian randomization studies with categorical exposures such as disease status or educational attainment should be interpreted in terms of liability, which may act via pathways through the categorical exposure and via independent pathways.

## Methods

### UK Biobank

We used data from UK Biobank, a large prospective cohort study of 503,325 individuals, aged between 38-73 years at baseline, who were recruited between 2006 and 2010 from across the United Kingdom. The vast majority of study participants were genotyped, completed questionnaire data at baseline and have linked records with secondary care data and other health registries. The cohort has been described in detail in previous publications, including information on genotyping ^53,54^.

For the PGS analyses, we used childhood phenotypes recalled by UK Biobank study participants in questionnaire data. Study participants were asked to describe their comparative body size at age 10 in relation to their peers (field ID 1687) as either “Thinner”, “Plumper” or “About Average”. We used this variable to construct an ordered categorical childhood BMI measure (Thinner 0, About Average 1, Plumper 2). Study participants were asked if they currently wear glasses (or contact lenses) (field ID: 2207) and if answering yes were asked the age that they started wearing them (field ID: 2217). We used these variables to create a binary variable reflecting glasses use at age 15. Individuals who reported not currently wearing glasses or reported starting use after the age of 15 were set to 0. Individuals who started wearing glasses at 15 or younger were set to 1. Study participants were asked their smoking status (field ID: 20116) and if they reported previous or current smoking were asked their age of smoking initiation (field IDs: 2867, 3436). We used these variables to create a binary variable reflecting smoking initiation at age 15. Individuals who reported being never smokers or who reported initiation after the age of 15 were set to 0. Individuals reporting smoking initiation at 15 or younger were set to 1.

For the ROSLA-L analysis, we used country of birth (within the UK) (field ID: 1647) and a measure of educational attainment based on self-reported age when leaving full-time education (field ID: 845) to define individuals who left school at aged 16 in England or Wales. A measure of lifetime smoking was generated using “pack years of smoking” (field ID: 20162), available for current or former smokers, with lifetime smoking set to zero for self-report never smokers (field ID: 20116). BMI was measured using height and weight measured during the initial assessment centre visit (field ID: 21001). Systolic blood pressure was measured using an Omron device (field ID: 4080) at study baseline. Townsend deprivation index (TDI) (field ID: 189) was measured at recruitment and is based on the participant’s postcode and regional measures of deprivation. A higher TDI score suggests higher levels of deprivation. Information on income (field ID: 738) was obtained at baseline using the touchscreen questionnaire. Participants were asked to report the category relating to their average total household before tax in British pounds (“Less than 18,000”, “18,000 to 30,999”, “31,000 to 51,999”, “52,000 to 100,000”, “Greater than 100,000”). For the purposes of this study, we created a binary variable (0 if individuals reported household income less than 18,000 and 1 if individuals reported any of the categories with an income greater than 18,000). Any individuals reporting “do not know” or “prefer not to answer” were set to missing. Glasses use was defined using the baseline questionnaire variable on whether individuals wear glasses (or contact lenses) (field ID: 2207).

We used summary data from a previous Genome-wide association study (GWAS) independent of UK Biobank to identify genetic variants putatively associated with educational attainment (P < 1×10^−5^) ^55^. Sets of independent variants were generated by LD clumping (R^2^ < 0.001, 10000 kb distance for clumps) the summary data in PLINK v1.9 ^56^. Weighted genetic risk scores were then constructed in UK Biobank study participants using GWAS summary data beta coefficients in PLINK v1.9 ^56^.

### Genetic analyses

Starting with the full sample of UK Biobank participants with genotype data, we restricted to individuals of “White British ancestry” as self-reported and verified by principal components analysis. We then removed closely related individuals identified using an in-house algorithm. The final sample included 337,006 individuals. More information on the internal quality control of UK Biobank data is contained in a previous publication ^57^.

We then calculated the association between the educational attainment PGS and the childhood phenotypes described above (BMI, glasses use, smoking initiation) using regression (logistic/linear) models adjusting for sex, age and the first 10 principal components.

As a sensitivity analysis, we also applied within-family models using a sample of 41,497 siblings from 19,588 families from UK Biobank. Siblings were identified using UK Biobank provided measures of IBS (kinship) and IBS0 (proportion of null loci) ^43^. Within-family models were as above but also included the family mean PGS (the mean PGS amongst all siblings in a family) as a covariate to account for parental genotypes ^43,44^. To account for collinearity between siblings, standard errors were clustered by family.

### Raising of the school leaving age analysis

For the purposes of the ROSLA analyses, we restricted the sample to individuals self-reporting that they were born in England or Wales. We then defined the pre-reform cohort as individuals born between September 1^st^, 1956 and August 30^th^, 1957 who self-reported leaving school at the age of 16. The post-reform cohort was similarly defined as individuals born between September 1^st^, 1957 and August 30^th^, 1958 self-reporting leaving school at 16. In the combined sample, we then evaluated differences between the groups using a linear (Education PGS, BMI, SBP, smoking, TDI) or logistic (income > £18,000, glasses use yes or no) model with a “reform” covariate (0 for pre-reform and 1 for post-reform). Heterogeneity p-values were defined as the regression p-value of the reform covariate.

Caveats with the ROSLA approach in UK Biobank, such as the correlation between birth month and self-report school leaving age, have been discussed previously ^27^.

### Data availability

This study used individual participant data from the UK Biobank with field IDs listed in the methods section. For details on accessing UK Biobank data please contact. All summary data are contained in the article or available upon reasonable request.

### Code availability

Statistical code for regression models is available on GitHub (https://github.com/LaurenceHowe/LiabilityScripts/blob/main/regression-models.R). Other queries can be addressed to the corresponding author.

## Acknowledgements

This research has been conducted using the UK Biobank Resource under Application Number 8786. Quality Control filtering of the UK Biobank data was conducted by R.Mitchell, G.Hemani, T.Dudding, L.Paternoster as described in the published protocol(doi:10.5523/bris.3074krb6t2frj29yh2b03×3wxj). The Medical Research Council (MRC) and the University of Bristol support the MRC Integrative Epidemiology Unit [MC_UU_00011/1]. NMD is supported by an Economics and Social Research Council (ESRC) Future Research Leaders grant [ES/N000757/1] and the Norwegian Research Council Grant number 295989. No funding body has influenced data collection, analysis or its interpretation. Thanks to Michel Nivard (VU), Ben Brumpton (NTNU) and Dan Benjamin (UCLA) for helpful comments on the coherency of the manuscript. This publication is the work of the authors, who serve as the guarantors for the contents of this paper.

## Author contributions

LJH conceptualised the project and performed statistical analyses. GDS proposed the ROSLA-L analyses. LJH and NMD drafted the first version of the manuscript. All authors contributed to interpretation of results and manuscript writing.

## Notes

### Competing Interest Statement

The authors have declared no competing interest.

### Author Declarations

This research has been conducted using the UK Biobank Resource under Application Number 8786.

